# SARS-CoV-2 VARIANT PREVALENCE ESTIMATION USING WASTEWATER SAMPLES

**DOI:** 10.1101/2023.01.13.23284507

**Authors:** I. López-de-Ullibarri, L. Tomás, N. Trigo-Tasende, B. Freire, M. Vaamonde, P. Gallego-García, I. Barbeito, J.A. Vallejo, J. Tarrío-Saavedra, P. Alvariño, E. Beade, N. Estévez, S. Rumbo-Feal, K. Conde-Pérez, L. de Chiara, I. Iglesias-Corrás, M. Poza, S. Ladra, D. Posada, R. Cao

**Affiliations:** Research Group MODES, Research Center for Information and Communication Technologies (CITIC), University of A Coruña (UDC), Campus de Elviña, 15071 A Coruña, Spain; CINBIO, Universidade de Vigo, 36310 Vigo, Spain; Galicia Sur Health Research Institute (IIS Galicia Sur), SERGAS-UVIGO, 36312 Vigo, Spain; Microbiology Research Group: meiGAbiome-Biomedical Research Institute (INIBIC)-Center for Advanced Research (CICA)-University of A Coruña (UDC)-CIBER of Infectious Diseases (CIBERINF), Servicio de Microbiología, 3^a^ planta, Edificio Sur, Hospital Universitario, As Xubias, 15006 A Coruña, Spain; University of A Coruña (UDC), Research Center for Information and Communication Technologies (CITIC), Database Laboratory, Campus de Elviña, 15071 A Coruña, Spain; Department of Biochemistry, Genetics, and Immunology, Universidade de Vigo, 36310 Vigo, Spain

## Abstract

The present work describes a statistical model to account for sequencing information of SARS-CoV-2 variants in wastewater samples. The model expresses the joint probability distribution of the number of genomic reads corresponding to mutations and non-mutations in every locus in terms of the variant proportions and the joint mutation distribution within every variant. Since the variant joint mutation distribution can be estimated using GISAID data, the only unknown parameters in the model are the variant proportions. These are estimated using maximum likelihood. The method is applied to monitor the evolution of variant proportions using genomic data coming from wastewater samples collected in A Coruña (NW Spain) in the period May 2021 – March 2022. Although the procedure is applied assuming independence among the number of reads along the genome, it is also extended to account for Markovian dependence of counts along loci in the aggregated information coming from wastewater samples.

## Motivation and background

During the last decade, wastewater-based epidemiological surveillance has emerged as a highly relevant discipline, with the potential to provide information by combining the use of analytical methods with the development of ad hoc modelling approaches. This surveillance has been widely used in recent years to accurately predict consumption patterns for numerous substances (EMCDDA, 2020). During the COVID-19 pandemic, processes for monitoring the viral load of SARS-CoV-2 in wastewater were developed for the first time in the Netherlands (Medema et al., 2020).

Around a third of the people primarily infected with SARS-CoV-2 in Spain were asymptomatic (Pollán et al., 2020). However, the percentage of asymptomatic cases depends on many factors, such as the average age and the degree of natural or artificial immunity in each population. In addition, a significant proportion of people infected with COVID-19, including symptomatic and asymptomatic, who were tested for fecal viral RNA tested positive from the initial steps of infection (Gupta et al., 2020) and tested positive persistently in rectal swabs even after nasopharyngeal testing was negative (Chen et al., 2020; Xing et al., 2020; Xu et al., 2020; Zhang et al., 2020; Cevik et al., 2021; Miura et al., 2021).

Due to all of the above, the genetic material of SARS-CoV-2 can be found in wastewater (Lodder and de Roda Husman, 2020), which has made the monitoring of the RNA viral load in wastewater an excellent tool for the epidemiological monitoring of the COVID-19 pandemic, as well as an efficient early warning method for the detection of outbreaks (Randazzo et al., 2020; Ahmed et al., 2020; Medema et al., 2020; Peccia et al., 2020; F Wu et al., 2020; Wurtzer et al., 2020). Likewise, the methods of massive sequencing of aggregate samples collected in wastewater treatment plants or in the sanitation network itself make it possible to obtain readings that include the mutations observed in the SARS-CoV-2 genome. With the help of appropriate statistical models and methods, estimates of the number of active cases of patients with COVID-19 can be obtained from the viral load quantification data at Wastewater Treatment Plants (WWTPs) (Vallejo et al. 2022).

On the other hand, as a result of the proliferation of SARS-CoV-2 variants, specific mutations have been monitored to study the evolution of variants (Bar-Or et al. 2021) and the total SARS-CoV-2 concentration (Radu et al. 2022). Recently, statistical methods have been proposed that make it possible to analyze the readings of mutation frequencies in the virus genome in order to obtain precise estimates of the proportions of variants (Barbeito et al. 2022, Gafurov et al. 2022, Karthikeyan et al. 2022, Radu et al. 2022, Valieris et al. 2022). In this paper, the joint mutation distribution is estimated using GISAID data and the variant proportions are estimated using maximum likelihood. The model can be formulated either assuming independence among the number of reads along the genome or allowing for Markovian dependence of counts along loci.

## Methodology

Since the genetic material of the samples collected at the WWTP is degraded as a consequence of the passage of wastewater through the sanitation network, the genomes collected are remarkably fragmented. On the other hand, each sample corresponds to the genetic material of the thousands of infected human beings among the almost 400,000 inhabitants of the metropolitan area of A Coruña. As a consequence of all this and of the amplicon technology used for massive sequencing (see Section 4), the available information corresponds to counts of mutation reads throughout a number of positions (loci) in the virus genome.

In the case in which clinical samples could be taken from individual patients, it would be possible to observe the complete RNA strand (or at least very large fragments of it that could be juxtaposed), which means having observations of the vector variable that considers which type of mutation has occurred at each locus. However, for the samples obtained at the WWTP, it is only possible to observe the frequencies of mutations in each of these loci in an aggregated manner on the set of individuals that have excreted that genetic material. As a consequence, the statistical methods for estimating the proportions of variants have to be designed for the data-generating process, aggregated, in individuals, and marginal, in loci, that occurs in this setup. We will now formulate this data-generating process.

A viral haplotype can be expressed as a vector *x* = (*x*_1_, …, *x*_*l*_), *l* being the number of genomic positions or loci. The set of feasible values for locus *x*_*i*_ is *A*_*i*_ = {0, …, *a*_*i*_}, where 0 refers to the reference allele and 1, …, *a*_*i*_ are indices identifying the alternative alleles (i.e. different types of mutations at locus *i*=1,…,*l*). As a consequence, *x* ∈ *H, H* being the Cartesian product. *A*_1_ × … × *A*_*l*_. We denote by *X* and *V*, respectively, discrete random variables modeling a haplotype and a viral variant sampled at random from the viral genomes in wastewater. For *r* viral variants *ν*_1_, …, *ν*_*r*_ the quantities 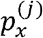, for *j* = 1, …, *r*, are defined as *P*(*X* = *x* | *V* = *ν*_*j*_). So 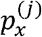, when *x* ∈ *H*, is just the haplotype distribution of variant *ν*_*j*_. By the total probability law, 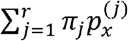, where *π*_*j*_ = *P* (*V* = *ν*_*j*_) is the unknown probability of the *j-*th variant. It is important to remark that, although the 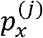 are also unknown, they can be estimated very easily without using the wastewater samples, e.g., from the viral genomes available at GISAID’s EpiCoV database.

If the viral genomic sequences could be fully observed in wastewater, the data would consist of a sample of haplotype vectors *X*_1_. …, *X*_*n*_. Given this “ideal sample” (not observable in wastewater, just for clinical patients), the observed sample can be modeled as follows. Consider, for each locus *k*, for *k* = 1, …, *l*, the probability *α*_*k*_ that the *k*-th locus of a viral genome selected at random is observed in the sample. The number of observations for locus *k* is 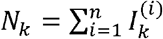, where 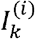 is a binary random variable indicating whether the *i*-th “ideally observed” haplotype has been actually observed at locus *k*. It is natural to model *N*_*k*_ as a random variable with binomial distribution, B(*n, α*_*k*_) being the expected number of reads at locus *k*. Its mean *n α*_*k*_ depends on the *α*_*k*_ probabilities, which are strongly determined by the sequencing technology and may greatly differ across loci. Since the are observable, in the following we condition on their observed values.

Given, for *k* =1, …, *l* and assuming that the sequencing technology does not affect the marginal distribution of *X*, it is possible to derive the distribution of the observed allele frequencies for each locus in the sample, *Y* = (*Y*_1_, …, *Y*_*l*_) where, for *k* = 1, …, *l*, 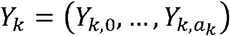 and 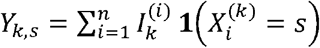. In the last expression, to avoid ambiguity, the superscript (*k*) is used to refer to the *k*-th component of *X*_*i*_, and **1** (*A*) is the indicator of event *A*. Clearly, 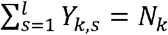, and, conditionally on *N*_*k*_, *Y*_*k*_ has multinomial distribution M(*N*_*k*_, qk) where 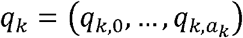, is a vector whose *S*-th component is 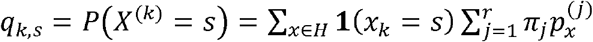.

Thus, the distribution of *Y* depends on the “known” haplotype probabilities within every viral variant estimated from available data 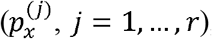, the number of reads at every locus (*N*_*k*_, *k* = 1, …, *l*), and the unknown variant probabilities (*π*_*j*_, *j* = 1, …, *r*) in the population of viral genomes sampled. The *π*_*j*_ can be estimated using available information and the observed allele frequencies in the wastewater sample. Assuming independence of the random variables, Yk, *k* = 1, …, *l*, and having observed the allele mutation frequencies collected in the vector *y* = (*y*_1_, …, y_*l*_) the likelihood (conditional on, *N*_*k*_, *k* = 1, …, *l*) is:

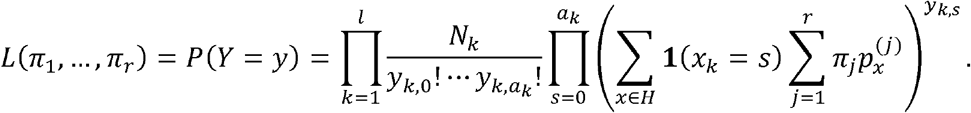

Maximum conditional likelihood estimates of (*π*_1_, …, *π*_*r*_) are obtained by maximizing *L* (*π*_1_, …, *π*_*r*_) constrained to *π*_1_ ≥ 0, …, *π*_*r*_, ≥ 0, 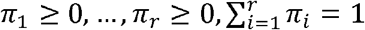e.g., using an augmented Lagrangian method.

### Markovian dependence among loci

The independence assumption among the random variables, *Yk, k* = 1, …, *l*, can be relaxed by just assuming a Markovian condition for the random vector *Y*:

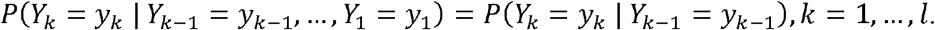

By assuming this condition, the likelihood becomes:

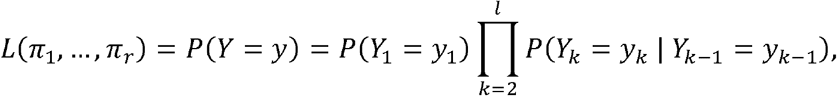

which just requires to deal with the conditional probabilities of the form *P*(*Y*_*k*_ = *y*_*k*_ | *Y*_*k*−1_ = *y*_*k*−1_), for *k* = 2, … *l*. Without loss of generality and for simplifying the notation, we consider *P(Y*_*2*_ = *y*_*2*_ | *Y*_*1*_ = *y*_*1*_) and assume that *a*_1_ = *a*_2_ = 1, i.e. just one type of possible mutation at loci *k* = 1,2. As a consequence, the joint distribution of (*Y*_1_,*Y*_2_) = (*Y*_1,0_,*Y*_1,1_,*Y*_2,0,_*Y*_2,1_) can be expressed in terms of the random vector *Z* = (*Z*_0,0_ *Z*_0,1_ *Z*_1,0_ *Z*_1,1_), where the random variable *Z*_*i,j*_ denotes the number of co-occurrences of mutation *i* in locus 1 and mutation *j* in locus 2. Indeed

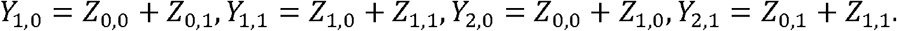

Now, since the random vector *Z* has a multinomial distribution: M(*N*_1,2,_(*p*_0,0_,p_0,1_,*p*_1,0_,*p*_1,1_)), where *N*_1,2_ is the number of joint reads at loci 1 and 2 and (*p*_0,0_,p_0,1_,*p*_1,0_,*p*_1,1_) is the vector with the probability mass corresponding to mutations (0 or 1) at loci 1 and 2, the joint probability mass of (*Y*_1_,*Y*_2_) is then straightforward:

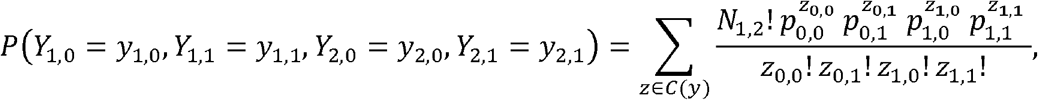

where z ∈ *C* (*y*) in the sum means that the values of z ranges over all possibilities such that *y*_1,0_ = *z*_0,0_ + *z*_0,1_, *y*_1,1_ = *z*_1,0_ + *z*_1,1_, *y*_2,0_ = *z*_0,0_ + *z*_1,0_, *y*_2,1_ = *z*_0,1_ + *y*_1,1_. The marginal probability mass of *Y*_1_ is even simpler:

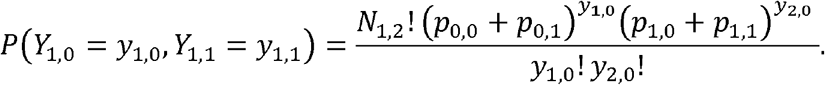

Using the definition of conditional probability, the conditional distribution becomes:

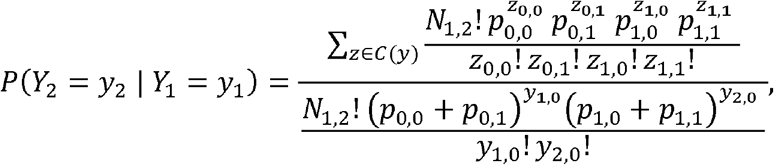

where the co-occurrence probabilities can be easily expressed in terms of the variants bivariate haplotype distributions, 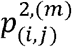, and the variant marginal distribution:

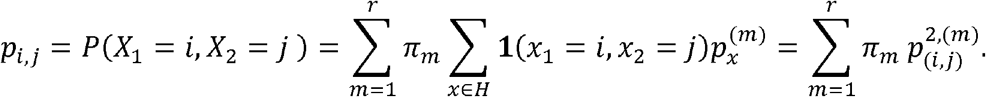

As a consequence, the likelihood in the Markovian dependence case can be written just in terms of the variants bivariate haplotype distributions and the unknown variant probabilities.

#### Simulations

Simulated data, as well as synthetic data coming from in vitro experiments, where the proportion of every variant is known, have been used to assess the quality of the method. We considered four scenarios.

Dataset #1 consists of simulated reads of 1 genome per variant without sequencing errors. The data were created from four different genomes from GISAID (consensus sequences), each genome corresponding to a different variant. A simulator of amplicon reads (with no sequencing errors) is applied based on the real coverage/depth profiles of ARCTIC protocol (obtained from real reads) and then those simulated reads are mixed in the percentages included in Table 1, which also contains the estimated percentages.

**Table 1:** Mixing variant percentages and their estimations for Dataset #1.

| Variant | B.1.1.7 | B.1.617.2 | B.1.621 | C.37 |
| --- | --- | --- | --- | --- |
| True percentages | 64% | 21% | 12% | 3% |
| Estimated percentages | 61.70% | 22.35% | 13.70% | 2.25% |

Dataset #2 also contains simulated reads without sequencing errors but of multiple genomes per variant. The data were created from four different genomes from GISAID (consensus sequences), each genome corresponding to a different variant. As for the previous dataset, a simulator of amplicon reads is applied based on the real coverage/depth profiles of ARCTIC protocol, obtained from real reads. The simulated reads are mixed in the percentages included in Table 2, which also contains the estimated percentages.

**Table 2:** Mixing variant percentages and their estimations for Dataset #2.

| Variant | B.1.1.318 | B.1.1.7 | B.1.351 | B.1.617.1 | B.1.617.2 | B.1.621 | C.37 |
| --- | --- | --- | --- | --- | --- | --- | --- |
| Real percentages | 0.00% | 53.00% | 0.00% | 0.00% | 27.00% | 7.00% | 13.00% |
| Estimated percentages | 0.95% | 52.17% | 0.91% | 1.17% | 24.88% | 7.05% | 12.86% |

Dataset #3 consists of mixing clinical samples created from real genomes reads obtained in the project EPICOVIGAL. For each variant, just one dataset is used and then the reads were mixed according to the percentages presented in Table 3. This table also includes the estimated percentages.

**Table 3:**
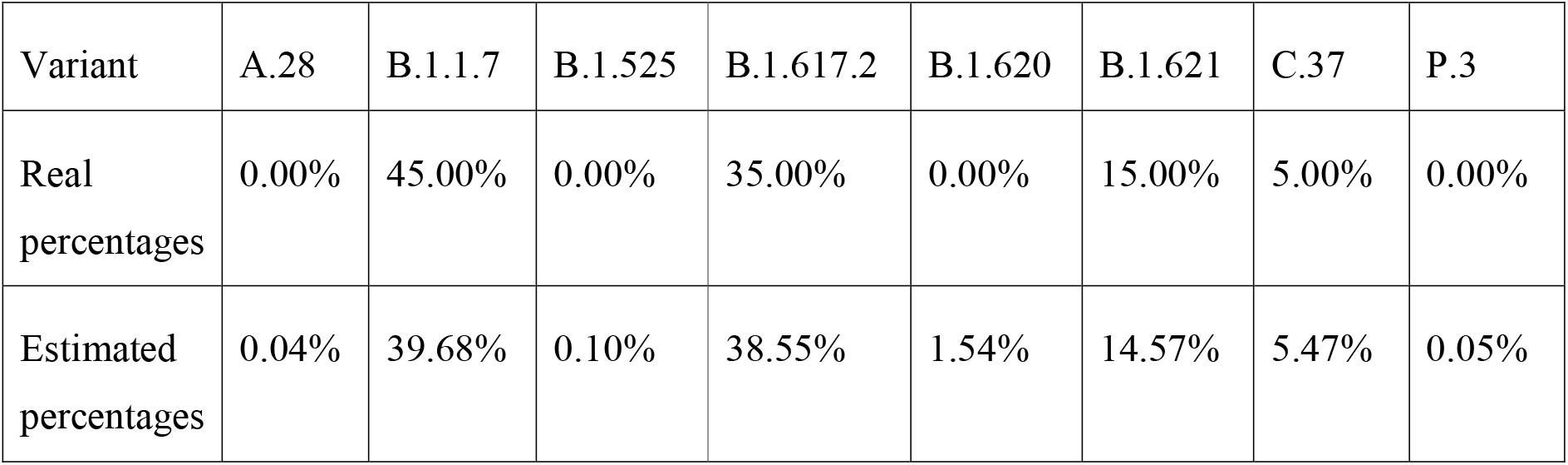
Mixing variant percentages and their estimations for Dataset #3.

Dataset #4 was also constructed by mixing clinical samples. It was created from real genomes reads obtained in the project EPICOVIGAL mixed in the percentages collected in Table 4, which also includes the estimated percentages.

**Table 4:** Mixing variant percentages and their estimations for Dataset #4.

| Variant | B.1.1.7 | B.1.351 | B.1.617.2 | P.1 |
| --- | --- | --- | --- | --- |
| Real percentages | 40.00% | 10.00% | 30.00% | 20.00% |
| Estimated percentages | 39.41% | 9.47% | 32.63% | 18.49% |

The results in Tables 1-4 show that the estimation error of the variant percentages is always below 2.7% for all the variants in Datasets #1, #2 and #4. For Dataset #3, the largest estimation error is around 5.3%. This happens for B.1.1.7, with a real percentage of 45%. This implies a relative estimation error of around 1/9.

#### Monitoring the evolution of variant proportions

The method presented is applied to monitoring the evolution of variant proportions using genomic data coming from weekly wastewater samples collected in A Coruña (NW Spain) in the period May 2021 – March 2022. This monitoring was part of the COVIDBENS project. It was an initiative carried out from April 2020 to March 2022 and financed by the public company WWTP Bens S.A., responsible for managing the WWTP in charge of purifying wastewater from the municipalities of A Coruña, Arteixo, Cambre, Culleredo and Oleiros, which comprise a population of nearly 400,000 inhabitants of the metropolitan area of A Coruña (NW Spain). The main objective of the project was to monitor the SARS-CoV-2 coronavirus epidemic in the metropolitan area of A Coruña.

COVIDBENS served as an early warning against possible outbreaks, since it proved to be able to anticipate between 2 and 3 weeks in the beginning of the pandemic waves with respect to the data on active cases reported by the health system (Trigo-Tasende et al. 2022). In addition, using the amount of genetic material of the virus present in the wastewater, nonparametric statistical models were used to estimate the number of infected people in the population (Vallejo et al. 2022).

Since December 2020, complying with the recommendation of the European Commission (https://ec.europa.eu/environment/pdf/water/recommendation_covid19_monitoring_wastewaters.pdf), the COVIDBENS team has been in charge of monitoring the emergence of new mutations and variants of SARS-CoV-2 in the wastewater arriving at the Bens WWTP using massive sequencing technologies. With the collaboration of Aguas de Galicia and EDAR Bens S.A., this challenge was tackled using two different strategies: 1) amplicon sequencing and 2) shotgun sequencing with enrichment of human respiratory viruses. The results obtained by the COVIDBENS team showed that both technologies are effective for the detection of SARS-CoV-2 mutations. Amplicon sequencing works very effectively to specifically detect SARS-CoV-2 mutations and variants, while shotgun sequencing should be oriented towards the epidemiological monitoring of respiratory viruses in general (SARS-CoV-2, influenza, RSV, etc.). It should be noted that these techniques made it possible to retrospectively detect mutations of the Alfa variant in samples from the metropolitan area of A Coruña at the beginning of December, a month before that variant was detected in clinical samples, demonstrating the great potential of genome analysis of SARS-CoV-2 in wastewater for early epidemiological detection of variants. Once the methodology was fine-tuned and contrasted, it was decided to implement amplicon sequencing as a routine mutation tracking method. The genetic material was extracted and sequenced from samples obtained weekly. Data were analysed for surveillance mutations recommended by ECDC (European Center for Disease Prevention and Control), guidelines updated on March 11, 2022 (https://www.ecdc.europa.eu/en/covid-19/variants-concern).

In the period May 2021 – March 2020, the SARS-CoV-2 sequencing work in wastewater carried out by COVIDBENS enabled reporting on the evolution in the presence of mutations and variants in the metropolitan area of A Coruña on a weekly basis. The data obtained through sequencing and analysis of mutations and variants of the virus can be viewed at the link http://www.edarbens.es/covid19.

The statistical methods presented in the second section were used to estimate weekly the proportions of SARS-CoV-2 variants in the metropolitan area of A Coruña. For facilitating visual interpretation, the estimates of the proportions along time were smoothed with a local polynomial regression estimator. The smoothing parameters were selected using plug-in methods (see Loader, 1999).

Figure 1 contains the smoothed estimates of the SARS-CoV-2 variant proportions along time in the period May 2021 – March 2022. The decrease of the Alpha variant (B.1.1.7) is shown at the beginning of the time period under study. The irruption of the Delta variant (B.1.617.2), its subsequent predominance and final vanishing are observed during this period. In the time interval December 2021 – January 2022, the Omicron variant (B.1.1.529) appeared and abruptly increased, which was parallel to a sudden decrease of the Delta variant. The BA.2 Omicron subvariant also exhibits a sudden increase in February 2022.

**Figure 1:**
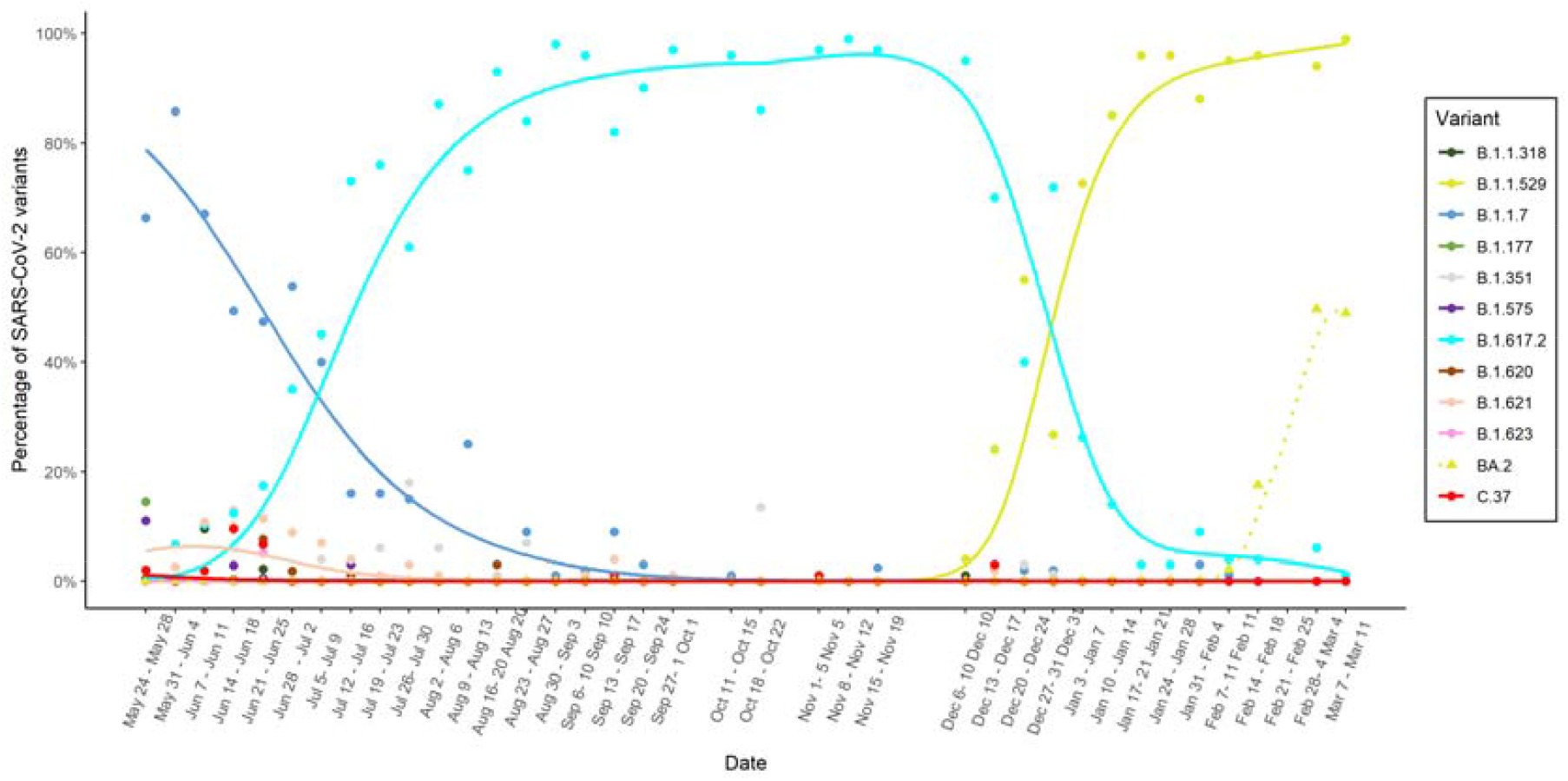
Smooth estimation of the SARS-CoV-2 variant proportions along time in the metropolitan area of A Coruña in the period May 2021 – March 2020.

## Data Availability

Data can be provided upon request.

## Notes

### Competing Interest Statement

The authors have declared no competing interest.

### Funding Statement

This work was supported by EDAR Bens S.A., A Coruna, Spain [grant references INV04020, 506 INV12120, INV05921 and INV148721 to MP], by the National Plan for Scientific Research, Development and Technological Innovation funded by the ISCIII, Spain - General Subdirection of Assessment and Promotion of the Research-European Regional Development Fund (FEDER) "A way of making Europe" [grant references PI15/00860 to GB, PI17/01482 and PI20/00413 to MP], by the GAIN (Xunta de Galicia, Spain) [grant references IN607A 2016/22 to GB, ED431C- 2016/015 and ED431C-2020/14 to RC, ED431C 2021/53 to SL and ED431G 2019/01 and COV20/00604 to RC and SL, by MINECO, Spain [grant references MTM2017-82724-R to RC], by the Spanish Network for Research in Infectious Diseases [REIPI RD16/0016/0006 to GB], by the "Innova Saude" Program, (INNOVAMICROLAB project) co-founded by the Galician Healthcare Service (SERGAS) and the Spanish Ministry of Science and Innovation, and by the Spanish Network of Research in Infectious Diseases (CIBERINFEC, ISCIII), and by the European Virus Archive Global (EVA-GLOBAL) project that has received funding from the European Union's Horizon 2020 research and innovation program under grant agreement No 871029. SR-F was financially supported by REIPI RD16/0016/006, KC-P by IN607A 2016/22 and the Spanish Association against Cancer (AECC) and JAV by IN607A 2016/22. DP was funded by grant EPICOVIGAL FONDO SUPERA-COVID19 from Banco Santander-CSIC-CRUE, Spain and grant CT850A-2 from ACIS SERGAS from the Conselleria de Sanidade of Xunta de Galicia, Spain.

